# The impact of early social distancing at COVID-19 Outbreak in the largest Metropolitan Area of Brazil

**DOI:** 10.1101/2020.04.06.20055103

**Authors:** Fabiana Ganem, Fabio Macedo Mendes, Silvano Barbosa de Oliveira, Victor Bertollo Gomes Porto, Wildo Navegantes de Araújo, Helder I. Nakaya, Fredi A. Diaz-Quijano, Julio Croda

## Abstract

We calculated the impact of early social distancing on the COVID-19 transmission in the São Paulo metropolitan area and forecasted the ICU beds needed to cope the epidemic demand by using an age-stratified SEIR model. Within 60 days, these measures would avoid 89,133 deaths.

---

The COVID-19 pandemic has led to the collapse of healthcare systems in several countries (1). The virus has a higher basic reproduction number (*R*0) (i.e., the average number of secondary cases generated by a primary case when introduced in a fully susceptible population) and case fatality rate (CFR) when compared with Influenza (*R*0:2,5-3,2 and CFR:0,4-2,9% versus *R*0:1,2-2,3 and CFR:0,15%-0,25%, respectively) (2–5). Among the confirmed cases in China, 18.5% were considered severe and 25.3% of those required intensive care (2). To tackle the spread of disease, a range of interventions have been implemented in China, including increasing test capacity, rapid isolation of suspected and confirmed cases and their contacts, social distancing measures, as well as restricting mobility (6).

In Brazil, the first confirmed COVID-19 case was reported on February 26th in the São Paulo city and, since March 16th, the state of São Paulo has recommended several social distancing measures. These include recommending that older adults and individuals with underlying chronic medical conditions stay at home as much as possible; cancelling mass events; reducing public transportation; closing schools, universities and workplaces; and maintaining only essential services.

The São Paulo Metropolitan Area (SPMA), one of the most populous urban area in the world, has 7,300 ICU beds registered in the National Council of Health Establishments (Cadastro Nacional de Estabelecimentos de Saúde, CNES: http://cnes.datasus.gov.br), 2,880 of which belong to the Public Healthcare system.

As the collapse of health care systems is the major concern for most countries hit by the pandemic, non-pharmacological interventions has been recommended to flatten the epidemic curve and gain time to prepare the health system to avoid shortage of ICU beds and healthcare workers needed to treat critically ill patients (4).

## The study

We evaluated the impact of early social distancing measures in the transmission of COVID-19 in the SPMA, and forecast the number of ICU beds necessary for COVID-19 patients in Brazil.

In 2009, the MoH established a mandatory notification for hospitalized cases of severe acute respiratory illness (SARI) through the National Disease Notification System (SIVEP-GRIPE). We retrieved all SARI cases reported on the SIVEP-GRIPE system between 26^th^ February and 30^th^ March. Those cases were included regardless of COVID19 confirmation as a proxy of COVID19 case. This proxy was used to calculate the time dependent reproductive number R(t) of confirmed COVID19 cases and was chosen in order to minimize the impact of shortage of RT-PCR tests. In addition, we also calculated R(t) using the daily number of COVID19 confirmed cases from the São Paulo state epidemiological records.

We calculated the R(t) during one month period in the SPMA, both, for COVID19 confirmed cases, as well as for SARI cases, and estimated the expected number of SARI cases requiring an ICU bed. The reproductive number at the beginning of the epidemic (R0) and during the epidemic (Rt) were calculated using the package R0 R Studio (3). The expected ICU demand was calculated using an age stratified SEIR model (7), which includes compartments for individuals requiring hospitalization and intensive care. The model parameters are described in Table 1.

**Table 1.**
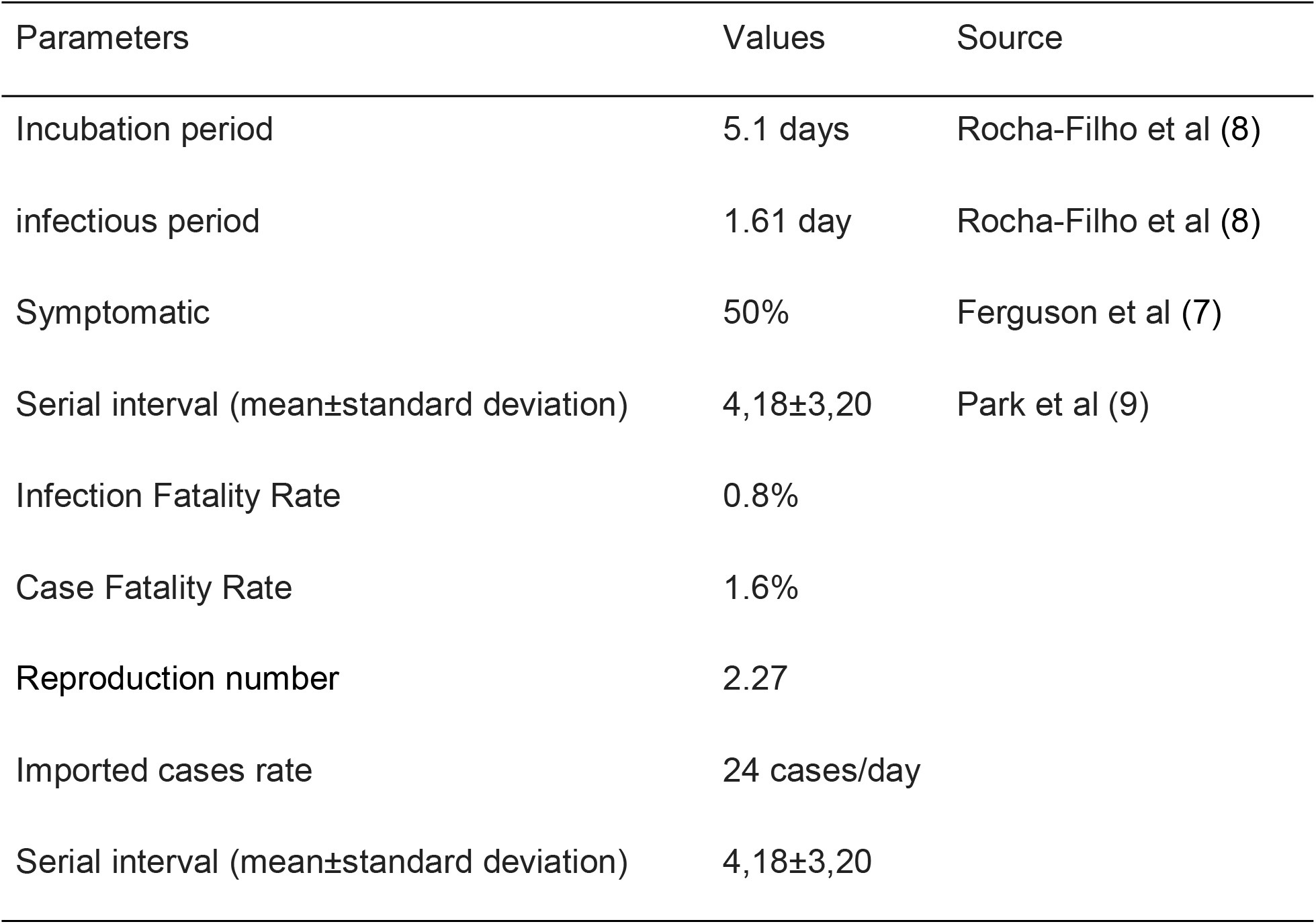
Parameters used in the age stratified SEIR model to forecast the ICU beds

Considering only the confirmed cases reported by São Paulo state, the R(t) was close to 2 throughout the assessed period with a large confidence interval. (Figure 1a). Underreporting and lack of confirmatory tests for COVID19 could directly affect these R(t) estimations. (Figure 1a). By analyzing the number of SARI available in the Epidemiological Surveillance of Influenza System (SIVEP-GRIPE), we found that the social distancing measures reduced the R(t) below to 1 with a more accurate confidence interval (Figure 1b). The R0 was used in the SEIR model to forecast the ICU beds needed and the number of deaths in a scenario without social distancing measures. The R(t) of SARI cases was used to forecast the scenario with social distancing measures.

**Figure 1.**
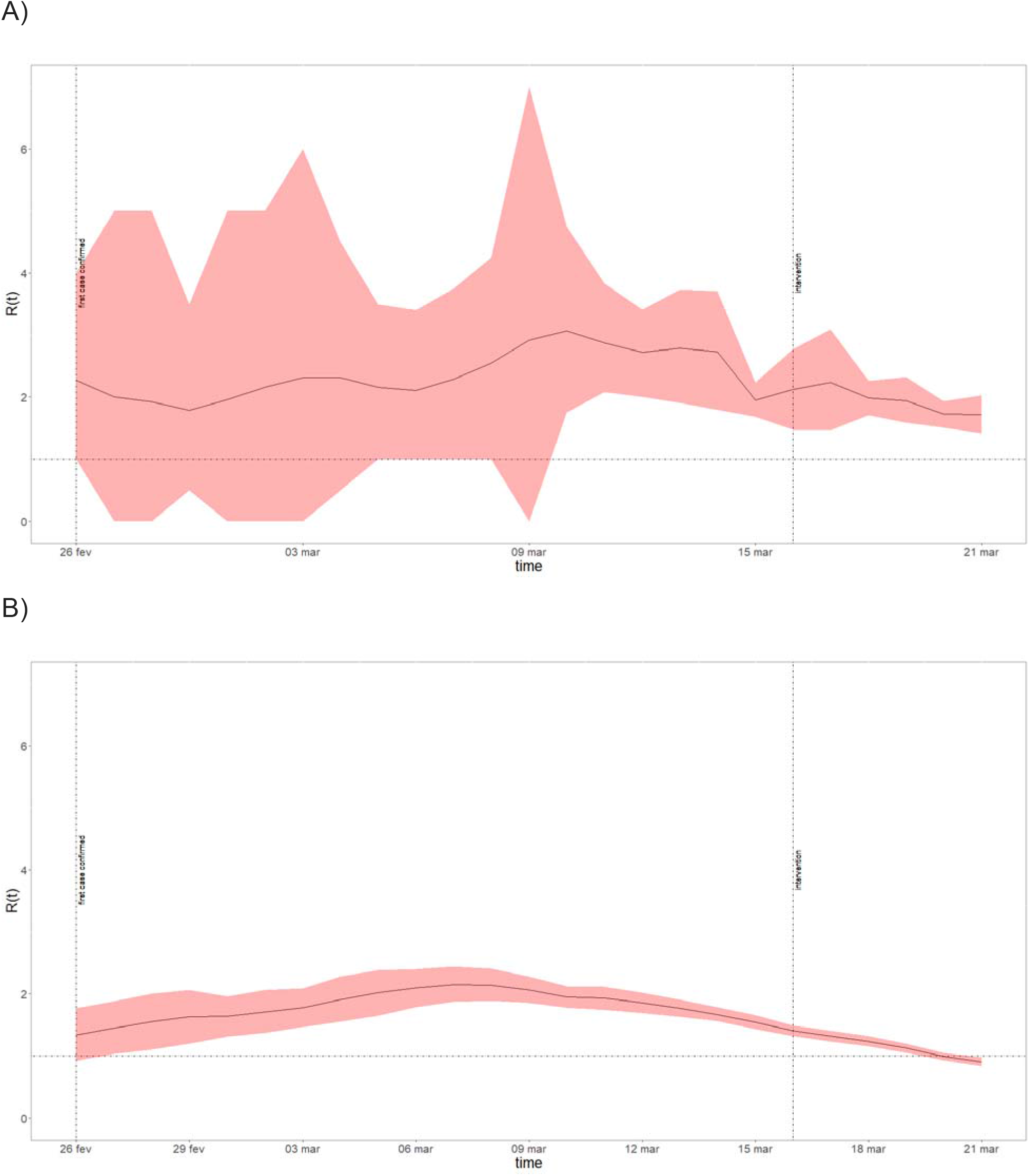
Time-dependent reproductive number R(t) estimated for the confirmed COVID19 cases (a), and for the SARI cases (b). Solid line corresponds to R(t), the red area of the confidence interval; the vertical dotted line represents, respectively, the date of the onset of symptoms of the first confirmed case in Brazil and date of the first social distancing measure implemented by the São Paulo state.

We defined the ICU bed capacity for COVID-19 as 20% of the total number of ICU beds in the SPMA. In the absence of social distancing measures, the model predicts that after 30 days, COVID19 patients would demand 5,384 ICU beds, which surpasses the current ICU capacity in 130%; furthermore, in the second month the ICU bed demand would be 14 times the ICU capacity. Overall, this would result in 1,783 deaths in the first month and 89,349 in the second month. While social distancing measures are maintained, the model predicts 317 deaths in the first month and a total of 1682 in the second. This scenario does not overburden the healthcare system which represents a maximum of 76% ICU beds capacity.

Using the severe cases notification systems, we identified that the social distancing measures implemented in the SPMA reduced the COVID19 R(t) to less than 1. If a similar level of social distance is maintained in April and May, during influenza seasonality, no additional ICU beds for COVID-19 patients will be needed in the SPMA.

We observed that the downward trend in hospitalized cases started on 9th March, before the intervention. This could be explained by a decrease in mobility documented since the first days of March in places like national parks, dog parks, plazas, and public gardens. According to a COVID-19 Community Mobility Report informed by Google this reduction was intensified and spread to other settings since the local government declared a state of emergency(8). This report suggested that intervention was effectively applied, and it is consistent with the transmissibility reduction observed. Furthermore, uor study indicated that social distancing measures impact should be monitored on daily based using hospitalized SARI cases, especially during the shortage of the COVID-19 confirmatory tests.

The baseline scenario shows that completely relaxing social distancing results in thousands of additional deaths. Figure 2 shows that the rate of fatalities per day increases dramatically when ICU capacity is overloaded. The simulation shows a swift change in the number of deaths per day in this scenario quickly spikes from 100, a week prior the system is overloaded, to a peak of 4,160 deaths per day in less than a month.

**Figure 2.**
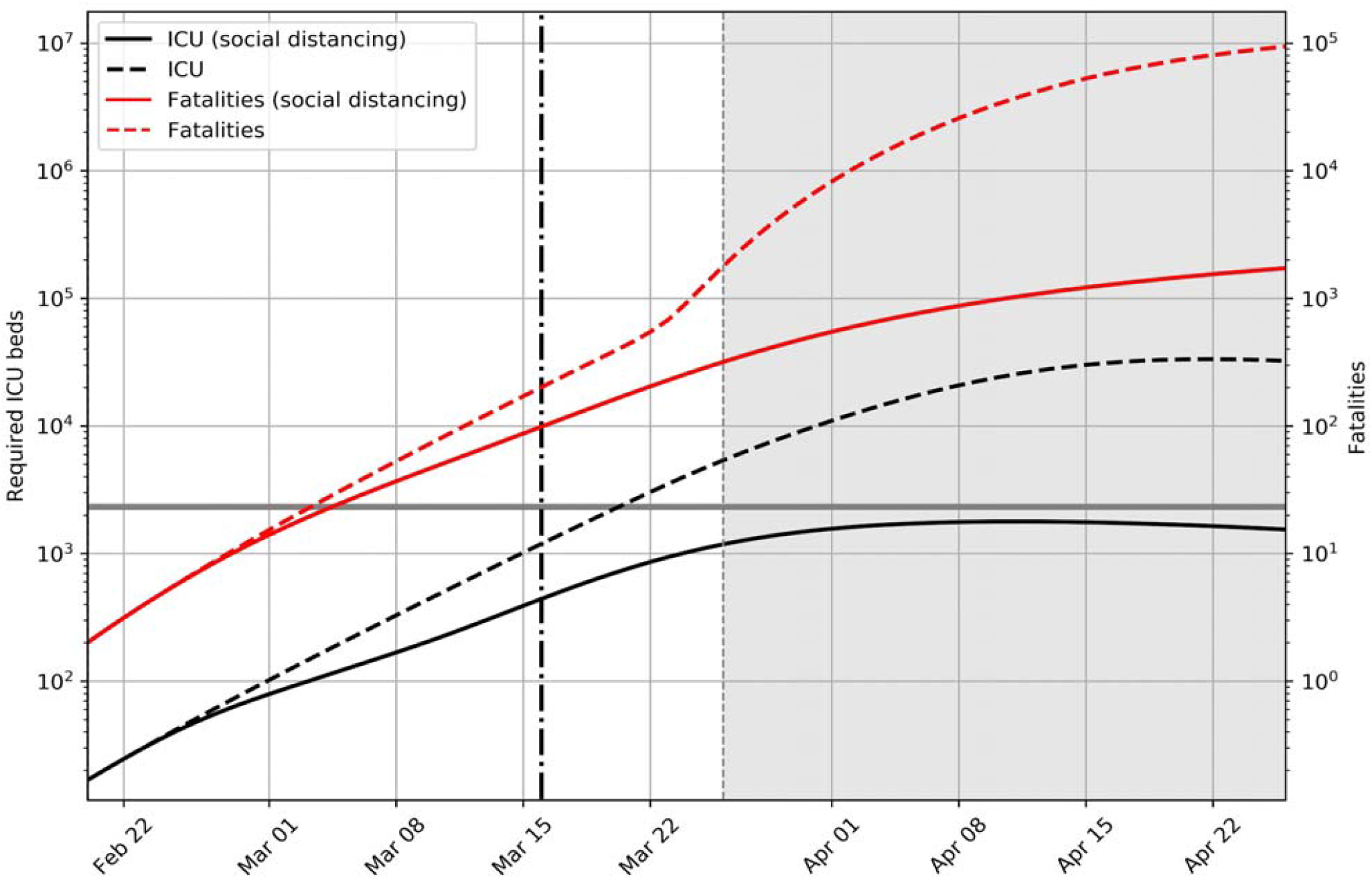
Estimation of the number of ICU patients (Black line) and fatalities (Red line). The solid line uses the time dependent reproductive number R(t) measured for SPMA and the dashed line represents the scenario in which no social distancing takes place. The vertical dashed line represents the date in which the local government declared a state of emergency and the horizontal gray line represents the number of available ICU beds.

A potential limitation is that many COVID-19 patients from nearby cities usually sought for medical care in the SPMA area, which is a reference for the state of São Paulo, leading to overburden of the healthcare system even if the epidemic is controlled in the SPMA. The last week of data has not been included to minimize the impact of delay in reporting.

## Conclusions

Despite the limitations, by using SARI electronic notification systems as a proxy for severe cases of COVID19 we reported a substantial decrease on the R(t) after two week of the implemented of social distance measures in the SPMA. These measures are expected to avoid 89,133 deaths within 60 days even without expanding the ICU bed capacity.

Fabiana S.G. dos Santos is a biologist and PhD candidate. Her main research interest is epidemiology of infectious diseases and mathematical modelling

## Data Availability

All data in this manuscript, including tables and files, are available.

## Acknowledgments

FADQ and JC were granted a fellowship for research productivity from the Brazilian National Council for Scientific and Technological Development – CNPq, process/contract identification: 312656/2019-0 and 310551/2018-8, respectively.

Dra. Maria Almiron, PAHO - Pan American Health Organization, Brazil Country Office for review the manuscript

## Notes

### Competing Interest Statement

The authors have declared no competing interest.

